# Time on Task in Psychotherapeutic/Psychoeducational Intervention with Intermittent Explosive Disorder

**DOI:** 10.1101/2024.09.12.24312716

**Authors:** Joseph Strayhorn, Stephen V. Faraone, Yanli Zhang-James

**Author notes:** Corresponding author: Yanli Zhang-James, MD PhD, Associate Professor, Department of Psychiatry and Behavioral Sciences, Norton College of Medicine, SUNY Upstate Medical University, 3732B NRB, 505 Irving Ave., Syracuse, NY 13210.

## Abstract

**Objective:** Anger control has been seen as a set of learnable skills. How much time is necessary for such learning? Comparisons with time requirements for other skills make it plausible that for many people, learning anger control may require well over 100 hours of time on task. Research interventions have been shorter -- a mean of 9 sessions was reported in one meta-analysis. In this study, our goal was to examine how much psychotherapeutic intervention is being delivered in the “real world” to patients with Intermittent Explosive Disorder.

**Method:** We studied a de-identified electronic health record data from TriNetX, collected from 87 medical institutions. We studied 32,322 individuals with Intermittent Explosive Disorder. We examined the distribution of the number of individuals across numbers of sessions received.

**Results:** The distribution for the numbers of sessions is highly skewed, resembling a curve of inverse proportion, or a Pareto function. The mode and the median were zero. Only about 25% of patients received any psychotherapy. For that subset, the median was 5 sessions, and the mean was 16. Approximately 10% received 9 visits or more; 5% 30 or more; 2% 50 or more. A large fraction of the psychotherapeutic labor was devoted to a small fraction of the patients: 80% of the sessions went to 7.5% of the patients.

**Conclusions:** The ability of health care systems to reduce the societal problem of aggression, at least by psychotherapeutic intervention, appears limited by the factors leading to low, or no, time on task.

## Introduction

Aggression, violence, maladaptive anger, and “man’s inhumanity to man” are perhaps the foremost problem of the human species. Psychotherapeutic and psychoeducational methods for helping people become less aggressive have received much study.

How long should we expect it to take, through new learning, to remove maladaptive aggressive habits and substitute for them new habits of kinder and more reasonable patterns? “Time on task” or “engaged time” is a major topic in the education literature (1, 2), but it is infrequently addressed in the mental health literature.

The definitive way to determine necessary hours would be a mastery learning approach: to deliver training to large numbers of people until sustained remission from maladaptive aggression is reached, and examine the distribution for the time on task required. Such a strategy has never been employed. Short of this, we can make guesses based on the complexity of the skills involved and the time required for learning other complex skills. We can also examine the times on task chosen for research interventions.

These approaches prompt a third question: In the “real world” of clinical practice, how much intervention are people with aggressive problems actually receiving? Is the typical amount of time on task enough for the health care system to reduce the societal burden posed by aggression? The current study addresses this.

### Estimating anti-aggression practice time from that required for other tasks

How long does it take to learn various skills? Of course, this varies widely with the learner, the teacher, the skill, the practice technique, and other factors. But here are some estimates people have made:

1. To become truly expert in playing the violin takes an average of about 10,000 hours. (3-5).
2. To become a concert pianist: aspiring students often devote about 1,400 hours per year during the teen years (6).
3. To be a competitive high school swimmer: “All but one or two of our subjects were swimming four hours (or more) a day, six and sometimes seven times a week. During the summer even more time was spent in practice.” This translates to at least 1250 hours per year. (6)
4. To develop expertise in mathematics, tennis, and research in neurology required similar time on task to that of pianists and swimmers, according to Bloom, 1985.
5. For an English-speaking adult to learn “general professional proficiency” in Spanish takes 600 to 750 class hours(7).
6. For an English-speaking adult to learn Japanese or Mandarin takes 2200 class(7).
7. The hours spent by the average US K-12 student in school were “6.87 hours per day and 178.71 days per school year, on average, for a total of 1,227 hours per year.” (8).
8. Required practice driving, New York State, for those with learners’ permits before taking a road test was 50 hours (9).
9. Amount to be spent on a 3 credit college course is 135 hours (10).
10. An expert in the videogame, *Call of Duty*, estimated that the time for an experienced videogame player to “get good at” *Call of Duty* is 50 to 100 hours (11).

How difficult are the skills of anger control? The answer to this varies extremely widely, depending upon the baseline level of skills in the learner. Hopefully, most of the population functions at passable levels with no “formal” anger control training. How much influence, for better or for worse, most people receive from the influences provided by parents, peers, teachers, religious teachers, and the entertainment media is impossible to count. The extent of trauma, brain injury, intellectual functioning, genetic influences, and reinforcement history all influence on how thoroughly ingrained habits of impulsive aggression are, and thus, how much time on task is required for change. If habits that are stable over time are more resistant to change, aggression appears to be one of those: a study measuring anger of children longitudinally in first, third, and fifth grade reported that “effect sizes for anger stability were substantial, with stability correlations for consecutive assessments ranged from 0.55 to 0.70.” (12)

The skill set relevant to nonaggressive functioning is large and complex. The treatment manual for Aggression Replacement Training, a widely used program (13) lists a “skillstreaming checklist” comprising 50 skills, for example answering a complaint, being a good sport, responding to failure, dealing with an accusation, and dealing with group pressure. For each of the 50 skills, the manual lists three to five steps in execution of the skill. It is very plausible that for some learners, practicing for one hour one each step is just the beginning of attaining mastery; this would necessitate about 150 hours of time on task.

Especially for children, since low reading skill can make school a daily source of frustration, reading is an anti-aggression skill (14-18). Patterson and colleagues reported that an average of 33 hours of one-to-one work resulted in an improvement of one grade equivalent.

A sampling of manuals on anger control skills (17, 19-23) reveals many component skills that could be included in psychoeducation or psychotherapy for aggression. Supplementary Table 1 lists some of the tasks for anger control. This set is not complete, but just a sampling of the range of items on the anger control to do list.

Examining the number of hours spent in other pursuits renders it plausible that certain learners would need hours on task numbering over 100, or in the hundreds, or even in the thousands, to achieve mastery in the skills of anger control and nonviolence. And given the often-disastrous life consequences of very poor levels of such skills, the devotion of such hours, if necessary and successful, would be time well spent.

### Anti-aggression practice time in intervention studies

Researchers face constraints: the greater the training time, the greater is the dropout rate, the longer the comparison group goes without the presumably useful intervention, and the longer the study takes. In a meta-analysis of psychological treatments for anger (24), the mean number of treatment sessions was as 8.5 (SD 3.72), with range from 3 to 40. The number of sessions was positively correlated with effect size. This meta-analysis reported distribution of the number of sessions delivered over the examined studies are listed in Supplementary Table 2, highlighting that the time on task for most research interventions was less than that of 3 or 4 school days, and less than that of one day of videogame play for some players.

In the US, courts are mandating anger management training as a consequence for violent behavior or in lieu of incarceration. An organization designed to meet this demand (25) states that their “4-hour class contains 4 sessions and is designed to meet employers’ requirements. Our 8-hour program is specially created to cover courts’ and probations’ standards.”

Effect sizes are not a monotonic function of time on task. McCloskey et al. (2008) reported large effect sizes for a 12 session, 10 hour intervention; Larden et al. found no effect from a program which typically encompasses 45 hours. The first of these studied adult outpatients who voluntarily signed up for the intervention; the outcome measures were from self-report of the participants. The second was with adult convicted violent offenders who began training in the prison system; the outcome measure was violent recidivism rate. There is great heterogeneity in effect per unit time.

### Therapy time on task in “real world” settings

How much time do people spend learning psychological skills in actual clinical treatment, not research studies?

According to the US National Comorbidity Study Replication, of people meeting criteria for Intermittent Explosive Disorder, only 14% saw any mental health service provider (26). This was the lowest rate of service utilization of all the diagnostic categories studied. Of those who did see a mental health services provider, the median number of visits over 12 months was 3.5 sessions. The adolescent supplement of that study (27), reported that only 6.5% of adolescents with Intermittent Explosive Disorder for 12 months were treated for anger.

The current study reports more “real world” data regarding time on task for people with Intermittent Explosive Disorder.

## Method

Intermittent Explosive Disorder is certainly not the only DSM diagnosis where aggression, anger, irritability, or violence can be a prominent symptom; it is, however, the only one where impulsive aggression *must* be present. We tallied the number of psychotherapeutic sessions for individuals diagnosed with this disorder. Because the distributions were very similar for adults and minors, we combined the subsamples for the analyses reported here.

The TriNetX Research Network contained de-identified data from 117.7 million patients across 87 healthcare organizations (HCOs) globally at the time of the study (January 31, 2024). Among them, 33,547 patients had at least one diagnostic code for IED (ICD-10-CM F63.81, or ICD-9-CM 312.34 and 321.35). Because the data contains only de-identified patient medical records, the study was determined to be exempt by the SUNY Upstate Medical University Institutional Review Board.

We excluded patients with missing years of birth and those from healthcare organizations outside the US. We also excluded those who were diagnosed with IED or had their first healthcare encounter on or after Jan 1st, 2023 in order to allow sufficient time to engage in therapies. We excluded children with diagnosis recorded at 5 years of age or younger, since the DSM 5 requires attainment of age 6 for diagnosis. After these exclusions, 32,322 people remained in the sample diagnosed with Intermittent Explosive Disorder.

Psychotherapy sessions were identified using the Current Procedural Terminology (CPT®) codes 90832, 90833, 90834, 90836, 90837, 90838, 90839, 90840, 90845, 90846, 90847, 90849, and 90853. In the rare event where more than one code was entered on the same day, these were counted as one session. The psychotherapy sessions were counted for each patient. For each number of sessions, the total number of patients who had been seen that many times was tabulated.

## Results

### Sample characteristics

A total of 32,322 patients with IED diagnoses were included in the analysis. Of these, 71.4% were male, 25.2% were female, and 3.3% had unknown sex. The racial distribution was as follows: 64.5% were White, 14.7% were Black or African American, 1.1% were Asian, 0.6% were American Indian or Alaska Native, 0.4% were Native Hawaiian or other Pacific Islander, and 18.8% were of unknown or other races. Additionally, 10.1% of patients were Hispanic or Latino, 65.7% were non-Hispanic, and 24.2% had unknown ethnicity. The mean age of the patients was 35.4 years, with a standard deviation of 17.6 years. The mean age at IED diagnosis was 25.5 years, with a standard deviation of 17.1 years. At the time of their first visit, 60.8% of patients were 18 years or older, while 39.2% were under 18. Medical records spanned from 1970 to 2022.

### Summary Statistics

In the total sample, the median and the mode for psychotherapy visits were zero: 75% received no psychotherapy. The total number of psychotherapy visits was 129,482; the mean number for the whole sample was 4.0. Around 9% received as many as the 12 visits utilized in the McCloskey et al. (2008) study of psychotherapy for Intermittent Explosive Disorder.

If we look only at the 25% of the sample (8,108 people) who undertook psychotherapy at all, i.e. who took part in at least one session, the mode for the number of visits is 1. The median number of sessions was 5, and the mean was 16. About 16% of those receiving any therapy made 12 visits or more.

### The distribution of therapy sessions

Table 1 summarizes the number of people who had each number of sessions. It also displays cumulative percent frequencies for the number of people, and for the number of visits.

**Table 1.**
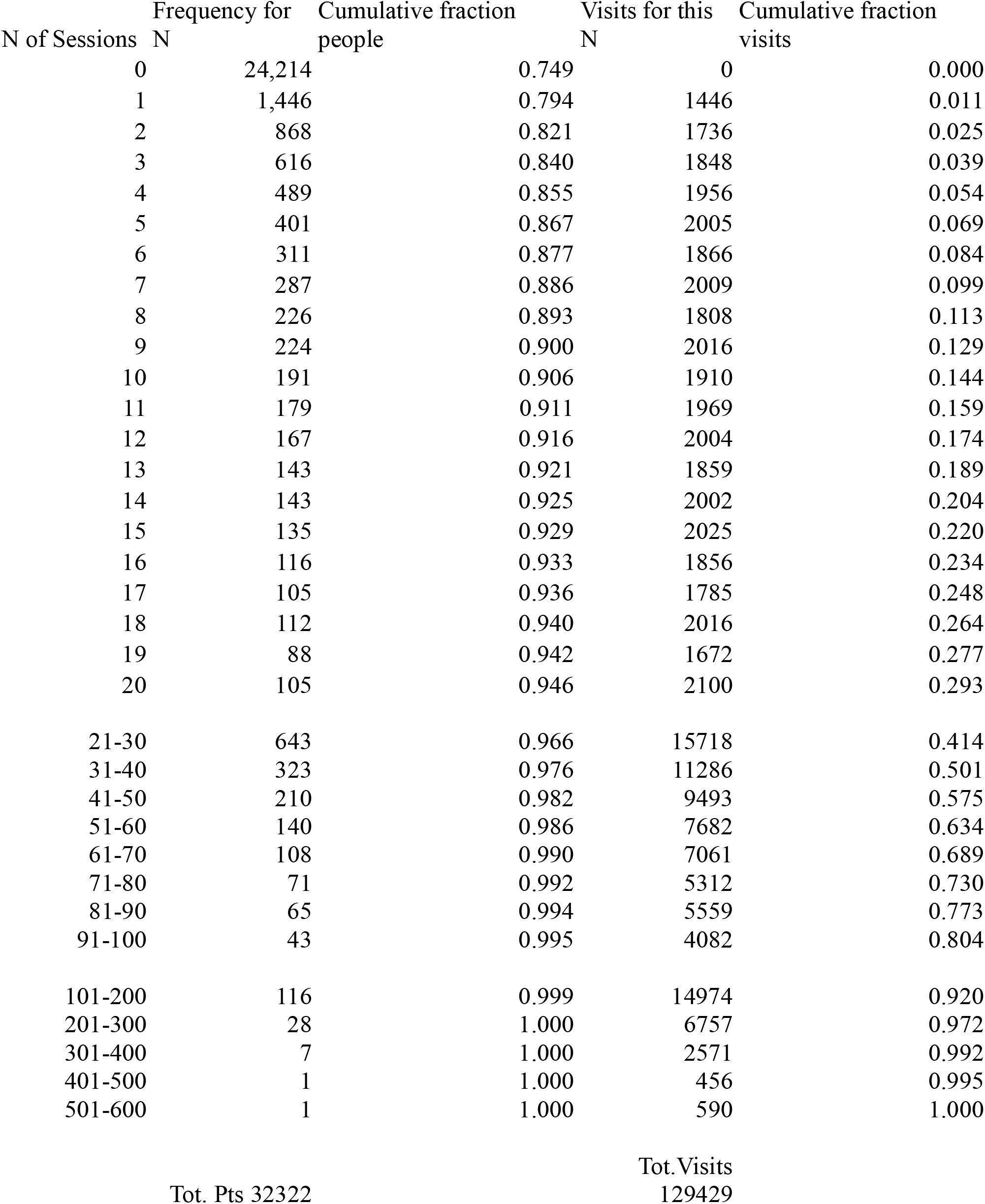
Distribution of Number of Sessions, Full Sample.

About 75% of the sample received no psychotherapy at all. About 90% received fewer than 10 sessions. Around 96% received fewer than the 30 sessions employed by Aggression Replacement Training. Around 98% had 50 sessions or fewer.

The shape of the distribution is illustrated by histograms. Because the frequencies past 80 sessions are too small to register on the graphs, the histograms are truncated at 80 sessions. Figure 1 displays the frequencies for the numbers of sessions.

**Figure 1.**
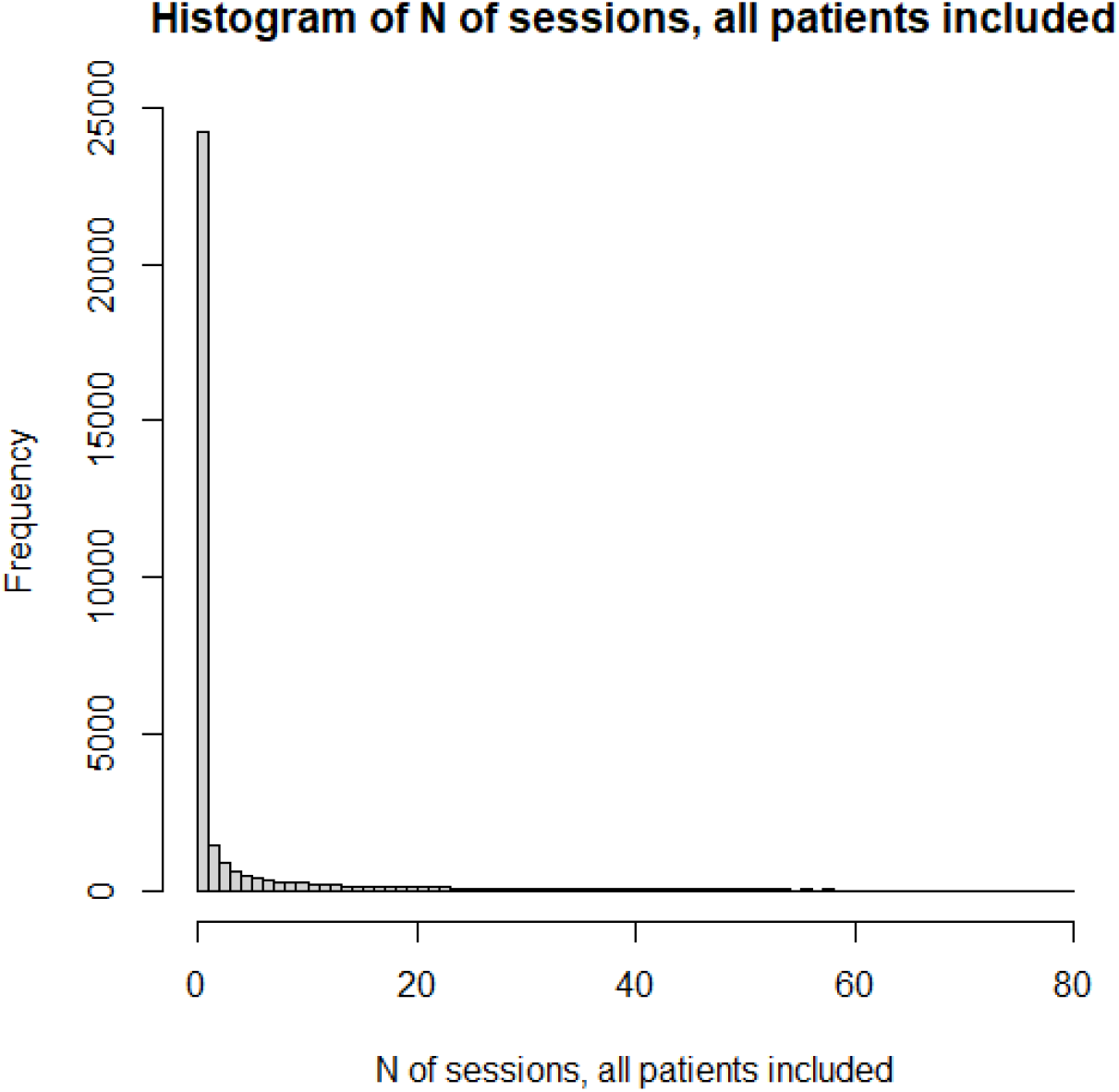

Because the number of “0” sessions is so large, we also present in figure 2 a histogram for the subset of patients who had at least one psychotherapy session.

**Figure 2.**
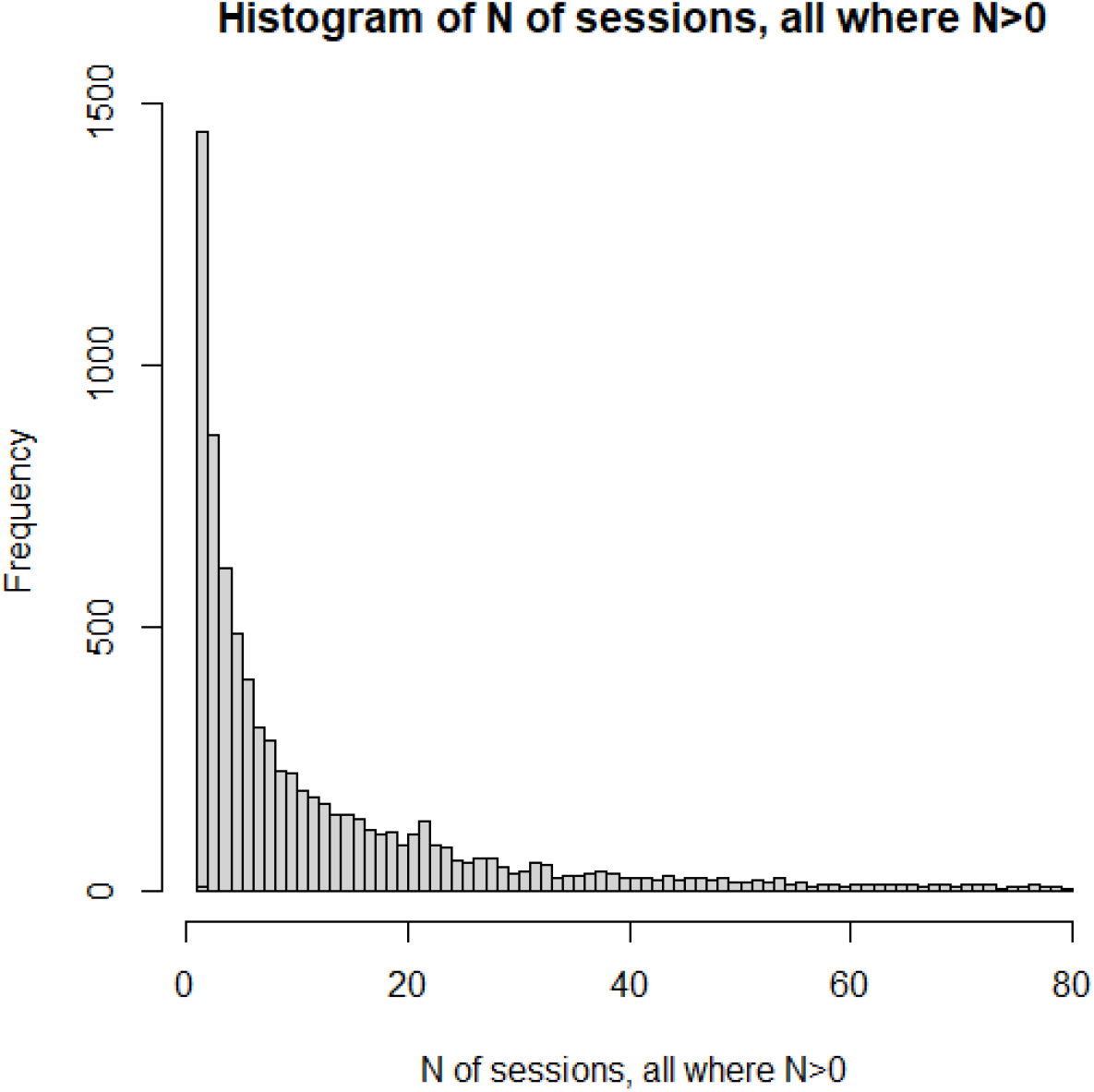

The distributions of number of psychotherapy sessions, as pictured in our histograms, do not even slightly resemble normal distributions. Rather, they resemble the shape of an inverse proportion. This is also the general shape of a form of the Pareto Distribution (28). The Pareto function, as well as the function of inverse proportion, yields linear results when the logarithm of the dependent variable is plotted versus the logarithm of the independent variable. Several analyses and graphs not included here supported the linearity of the log-log relation.

The graph of the cumulative fraction of the sample who came for a given number of visits is as in Figure 3.

**Figure 3.**
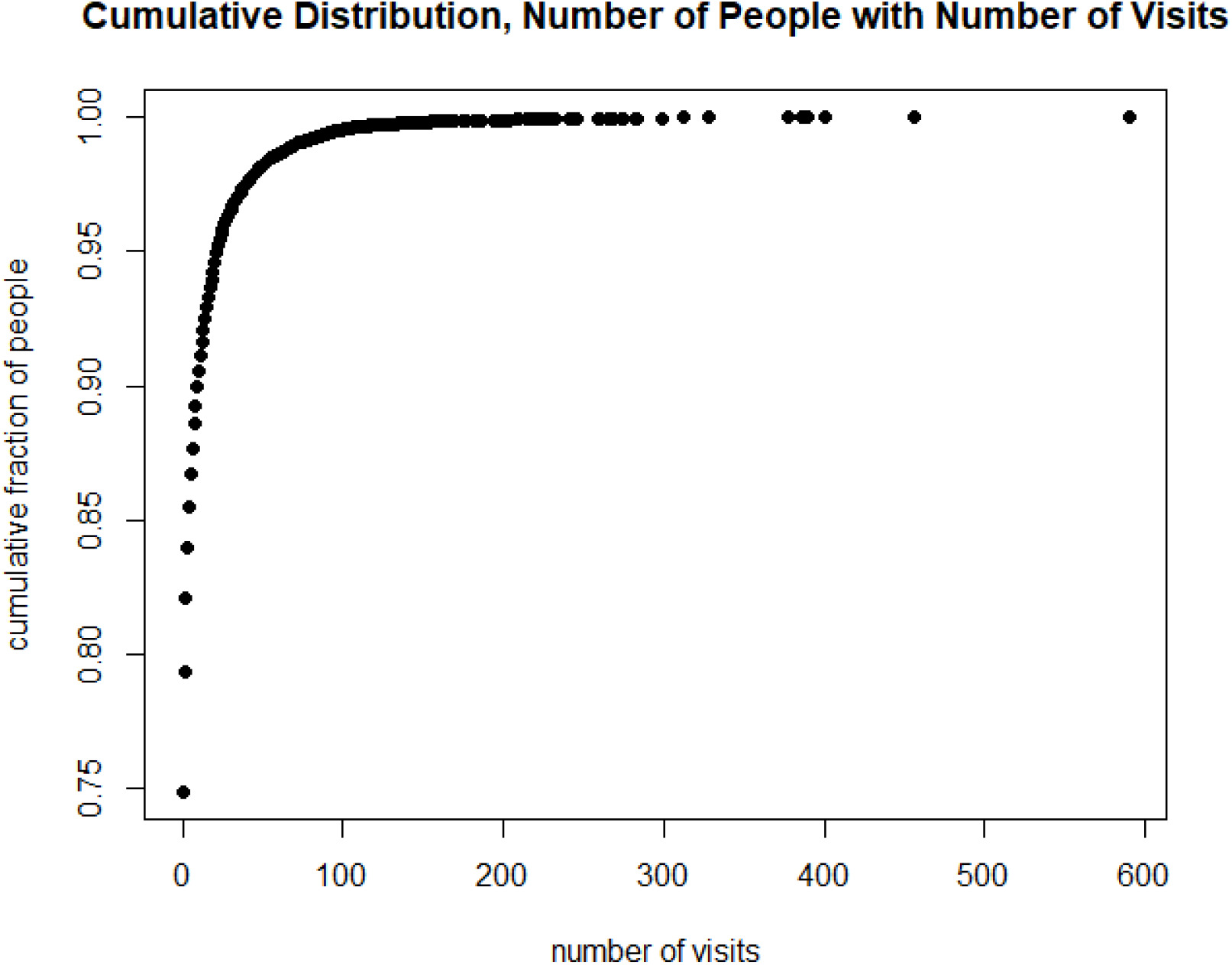

A plot of the cumulative fraction of individuals versus the cumulative fraction of visits for our total sample yields the curve in Figure 4; this sort of plot has been called the “Lorenz Curve” (29, 30). Such curves, generated with economic data, have produced very similarly shaped plots regarding income and wealth among a population; they have also been applied to distributions of botanical variables such as plant size and fecundity (31).

**Figure 4.**
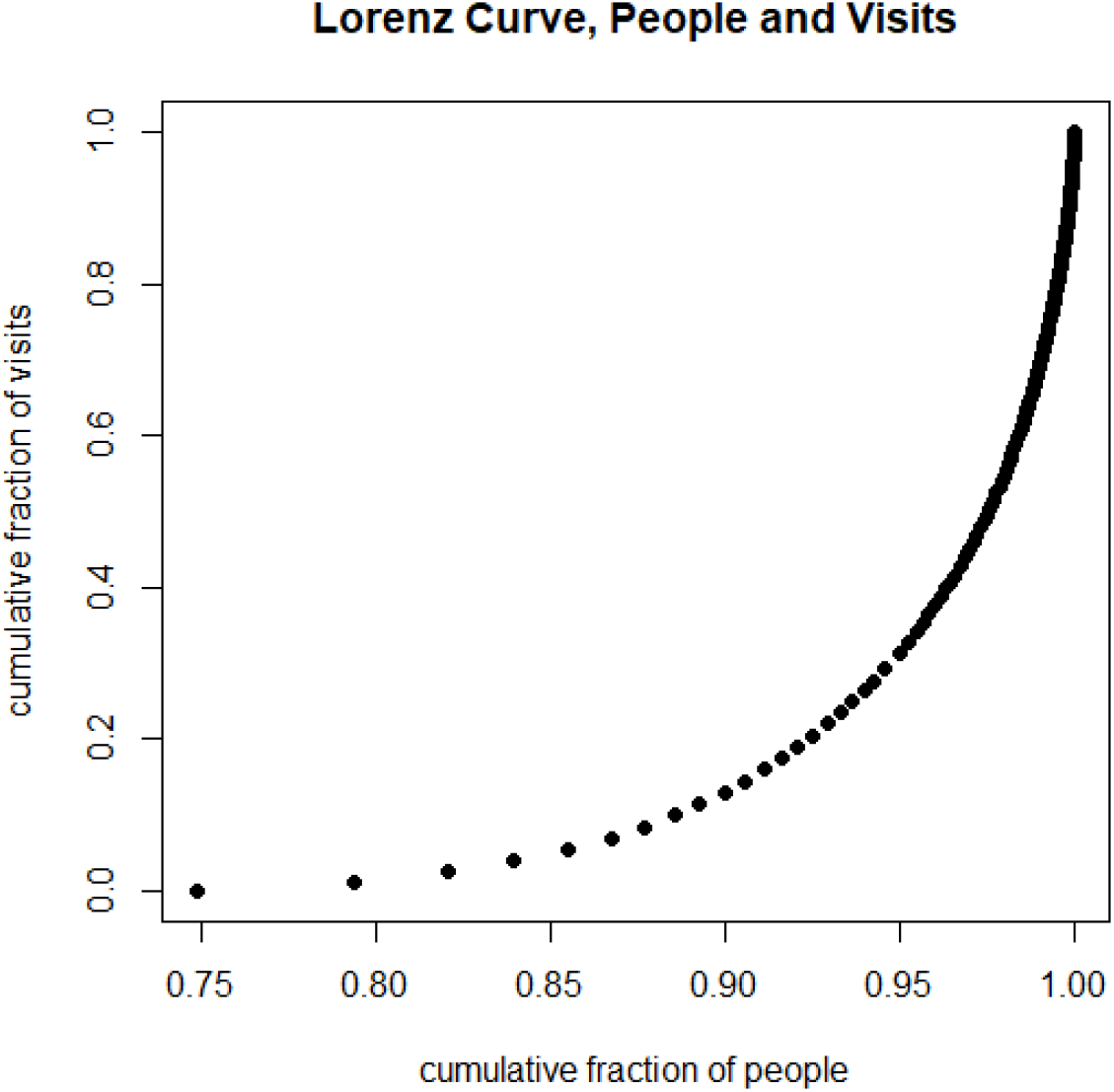

Figure 4 allows a determination of variants of the “80-20 rule.” As pictured in Figure 4, only about 20% of the total number of psychotherapy visits (129,429 visits) were accumulated by the first 92.5% of people. Thus (after subtracting both fractions from 1) 80% of the sessions were received by 7.5% of the people.

The visits we counted represent an upper limit that almost certainly overestimates the sessions actually devoted to the topic of aggression. In the same sample, Intermittent Explosive Disorder was found to be comorbid with almost all other psychiatric conditions (32). It is very likely that psychotherapy visits were devoted to anxiety, depression, and other comorbid conditions, in addition to those devoted to anger control.

## Discussion

Our results reveal two striking findings. First, the total number of sessions (and thus, time on task) in psychotherapy, for the vast majority of our sample, is far too low to produce useful results. Second, of the psychotherapeutic resources spent on this set of people, a large fraction of the labor is spent on a small subset of the people.

Let us call T1 the time spent necessary for skills of comparable complexity to anger control; T2, the time allocated in research interventions for psychotherapy of impulsive aggression, and T3, the median amount of time actually carried out in psychotherapy. Our examination of real world data and literature suggests that T1 > T2 > T3.

A great deal of work has been done in designing and testing psychotherapeutic interventions for aggressive behavior. There have been at least 21 meta-analyses of psychotherapy for aggression (33)! But for the vast majority of individuals diagnosed with Intermittent Explosive Disorder in our sample, all this work is irrelevant. For the vast majority, there was either no time or very little time devoted to psychotherapy.

The Pareto Distribution and some approximation of the 80-20 rule have been applied to numerous situations (34). The 80-20 rule is a consequence of inequality: for example, if each person had the same number of therapy visits, 80% of visits would be received by 80% of people. Our distribution of psychotherapy sessions, in which 80% of the sessions went to 7.5% of people, displays even greater inequality than the examples usually offered for the Pareto function.

A habit of impulsive aggression can be devastating to the life trajectory not only of the patient, but also of the recipients of the aggression. If the amount of time spent on a college course yielded much better results than much smaller time investments, the larger efforts would unquestionably be worthwhile; however, we know almost nothing about how much could be achieved by those levels of time on task or higher..

A limitation of this study is that we have no way of counting the amount of time on task that patients spent in homework between sessions. Although it is possible that some of them may have received a few hours of training which they supplemented by numerous hours of independent work at home, we are not optimistic enough to imagine that this takes place very frequently.

Another limitation is that psychotherapeutic work may have taken place during visits coded as evaluation and management or other non-psychotherapeutic codes. But since clinicians can code psychotherapy visits along with management codes if a significant time is devoted to it, it’s doubtful that there is a great deal of psychotherapeutic work missed.

A third possible limitation is that the people with an Intermittent Explosive Disorder diagnosis recorded in our sample represents only about 0.027% of the total pool of patients from which we drew, even after adjustment for the fraction eliminated from our sample. This represents a small fraction of the estimated prevalence of Intermittent Explosive Disorder in the population (35, 36). However, if this is because clinicians set a high bar for assigning the diagnosis, and have selected more aggressive individuals, we might guess that the investment of time on task could be even lower for those with lesser degrees of aggression. Our results on the sparsity of psychotherapeutic intervention are consistent with those of National Comorbidity surveys mentioned above, which gave an impression of even fewer psychotherapy sessions devoted to Intermittent Explosive Disorder than our results do.

What are the societal implications of these findings? We could exhort the health care system to seek ways of delivering more time on task to psychotherapy for aggression. But such exhortation could be in vain: it could be that the factors producing low time on task – perhaps finances, transportation, motivation, person-power, wish for quick results, devotion to the biomedical model? -- are deeply entrenched and may be as difficult to change as aggression itself. We don’t include calculations here, but if medical insurance programs had to pay for the number of psychotherapy hours really required for meaningful change, for all of the patients who could benefit, this would almost certainly “break the bank” of medical insurance, as well as requiring a vast increase in the mental health workforce.

Kazdin and Blase (37), who have examined at length the constraints on providing enough psychotherapy to relieve the societal burden of mental illness, spoke of the need for a “rebooting” of psychotherapy research and practice. They stated:

“Most people who might benefit from services for their dysfunction do not receive care. Additional resources in terms of person power might help. However, the dominant model of treatment delivery in clinical practice focuses on in-person treatment provided to individuals or relatively small units (groups, family, and couples). The model constrains the ability to reach individuals in need, even if the number of mental health professionals doubles.”(page 33)

One logical option is relying less on the health care system and more on other systems for psychological skills training. The education system is a candidate: arguably, the culture of education is more disposed to the investment of sustained hours of learning time. An argument against this is that many schools are not succeeding even in the most basic educational goal, that of teaching students to read (38). A counterargument is that aggression, anger, and violence are among the major reasons why some schools are not succeeding, and taking on fully the task of teaching the psychological skills thought to remedy those problems may be more successful than hoping that the health care system can do the job.

**Supplementary Table 1.**
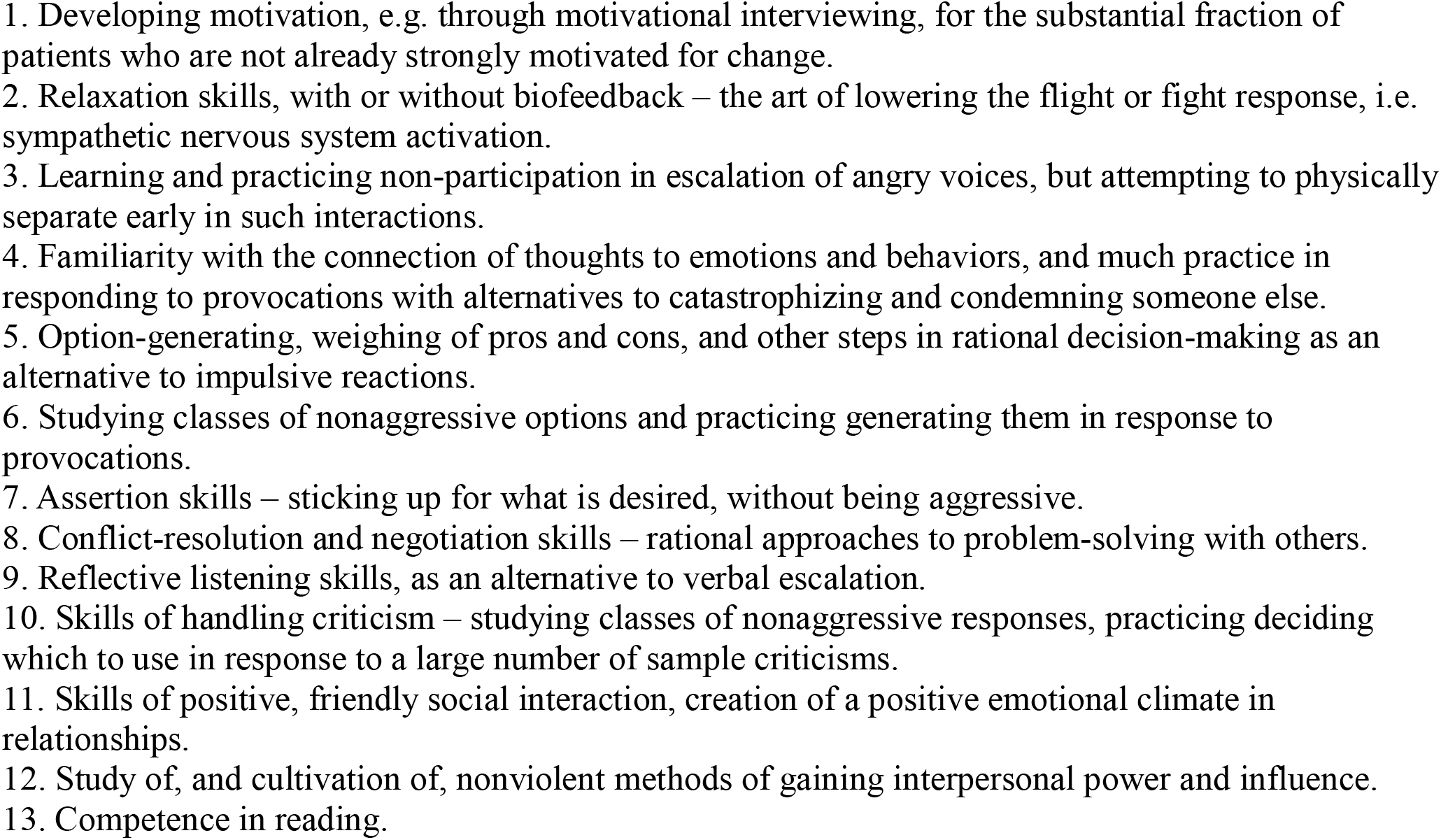
A sampling of skills or tasks on the to do list for anger control.

**Supplementary Table 2.**
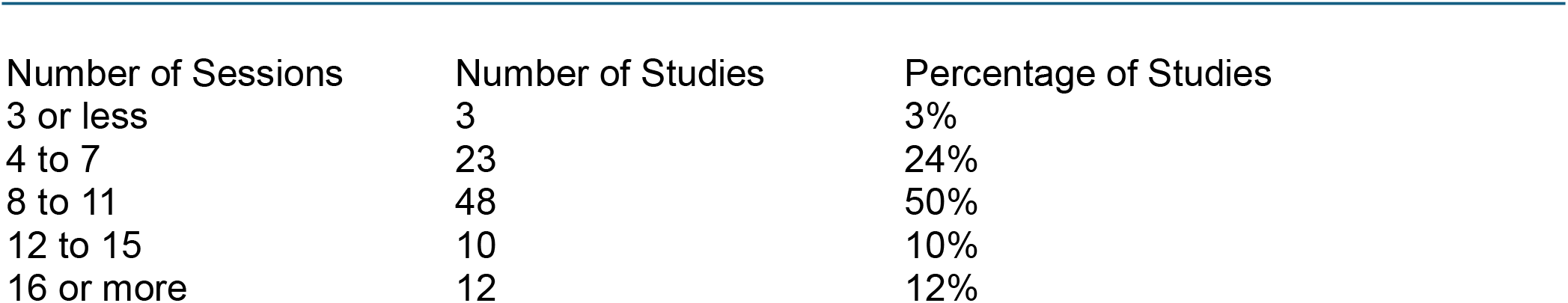
Distribution of Studies by Number of Sessions (Saini, 2009)

| Number of Sessions | Number of Studies | Percentage of Studies |
| --- | --- | --- |
| 3 or less | 3 | 3% |
| 4 to 7 | 23 | 24% |
| 8 to 11 | 48 | 50% |
| 12 to 15 | 10 | 10% |
| 16 or more | 12 | 12% |

## Data Availability

All data produced in the present work are contained in the manuscript, and available online at MedRxiv. De-identified medical records are only available directly from TriNetX.

